# An algorithm to identify patients with rare genetic disorders and its real-world data application

**DOI:** 10.1101/2023.01.27.23285056

**Authors:** Bryn D. Webb, Lisa Y. Lau, Despina Tsevdos, Ryan A. Shewcraft, David Corrigan, Lisong Shi, Seungwoo Lee, Jonathan Tyler, Shilong Li, Zichen Wang, Gustavo Stolovitzky, Lisa Edelmann, Rong Chen, Eric E. Schadt, Li Li

## Abstract

**Objectives:** Develop a digital phenotyping algorithm (PheIndex) using electronic medical records (EMR) data to identify children aged 0-3 who have been diagnosed with genetic disorders or present with illness with an increased risk for genetic disorders from a mother-child cohort.

**Methods:** We established 13 criteria for the algorithm where two metrics – a quantified score and a classification – were derived. The criteria and the classification were validated by chart review from a pediatrician and clinical geneticist. To demonstrate the utility of our algorithm in real-world evidence applications, we examined the association between size of carrier screening panel (small/≤4 genes [CS-S] vs large/≥100genes [CS-L]) undertaken by mothers prior to delivery, and children classified as presenting with illness with an increased risk for genetic disorders by our algorithm.

**Results:** The PheIndex algorithm identified 1,088 such children out of 93,154 live births and achieved 90% sensitivity, 97% specificity, and 94% accuracy by chart review. We found that children whose mothers received CS-L were less likely to be classified as presenting with illness with an increased risk for genetic disorders and a decreased need to have multiple specialist visits and multiple ER visits, compared to children whose mothers received CS-S.

**Conclusions:** The PheIndex algorithm can help identify when a rare genetic disorder may be present, and has the potential to improve healthcare delivery by alerting providers to consider ordering a diagnostic genetic test and/or referring a patient to a medical geneticist or other specialists.

**Article Summary:** Algorithm using EMR data to identify children who have been diagnosed with a genetic disorder or present with illness with increased risk of genetic disorders.

**What’s known on this subject:** With over 7000 Mendelian disorders, identifying children with a specific rare genetic disorder diagnosis through structured EMR data is challenging given incompleteness of records, inaccurate medical diagnosis coding, as well as heterogeneity in clinical symptoms and procedures for specific disorders.

**What this study adds:** We developed a digital phenotyping algorithm using electronic medical records (EMR) data to identify children aged 0-3 who have been diagnosed with genetic disorders or present with illness with an increased risk for genetic disorders from a mother-child cohort.

## Introduction

The widespread adoption of electronic medical record (EMR) systems has the potential to enable large-scale population-based studies characterizing patients with rare disorders.^1^ While identifying genomic information from EMR systems would assist in identifying such patient populations, with groups from Clinical Sequencing Exploratory Research (CSER) and Electronic Medical Records & Genomics © (eMERGE) representing such efforts, they have noted that genetic information is most commonly stored in unstructured formats such as PDF files or in paragraphs of free text, making genetic testing results difficult to locate.^2,3^ Additionally, CSER and eMERGE have not pursued a global approach to identifying patient populations with confirmed genetic disorders, or patients yet to be diagnosed with a genetic condition but rather whose medical records indicate that diagnostic genetic testing is warranted. Indeed, digital phenotyping studies using EMR data have largely focused on identifying populations with specific individual diseases, such as extracting patients with pediatric epilepsy, childhood obesity, or Noonan syndrome.^4-7^

When using EMR data to identify patient populations affected with rare genetic disorders, focusing on a specific rare genetic disorder diagnosis for any given patient is error-prone for many reasons. First, of 6519 rare disorders assessed, only 11% have International Classification of Disease 9 (ICD-9) codes and 21% have ICD-10 codes; some ICD codes are nonspecific, often with multiple phenotypes corresponding to a single ICD code.^8^ Furthermore, physicians and clinicians sometimes log certain ICD codes as they rule in or out a given condition, or when a condition is part of a differential diagnosis, yet still unconfirmed. Diagnosis codes may also be inaccurate or incomplete.^9^

Accordingly, algorithms that assess the risk of genetic disorders have the potential to improve healthcare delivery by assisting physicians and clinicians with clinical decision-making, including guiding when to order a diagnostic genetic test and/or refer a patient to a medical geneticist or other specialists may be indicated. Further, such algorithms could also be leveraged to identify rare genetic disorders patient populations to carry out cross-sectional and longitudinal epidemiological studies, assess healthcare utilization, and flag patients who may be considered for participation in specialized undiagnosed disease programs and precision medicine initiatives as underdiagnosis of rare genetic disorders is not uncommon.^10^

As a collaborative, multidisciplinary team, we developed a digital phenotyping algorithm that used structured EMR data and assessed 13 criteria to identify patients from birth to 3 years of age who have been diagnosed with a rare genetic disorder or who are at high risk for such a diagnosis. We tested our algorithm using a real-world dataset comprised of 93,154 live births with children linked to mothers’ medical records in a large academic health system. We validated the algorithm through blinded chart review by a pediatrician and a clinical geneticist.

To demonstrate the real-world evidence application of our algorithm, we examined the health outcomes of children whose mothers received carrier screening; specifically, whether there was an association between children who were classified as presenting with illness with an increased risk for genetic disorders by our algorithm, and the size of the carrier screening panel received by the mothers of these children. To the best of our knowledge, we are the first to generate a digital phenotyping algorithm beyond using ICD codes to identify children presenting with illness with an increased risk for genetic disorders and employed this algorithm to assess healthcare outcomes in a large, diverse, pediatric population.

## Methods

### Construction of mother-child cohort

We obtained de-identified EMR data through June 30, 2020 from the Mount Sinai Health System (MSHS). In total, we identified 93,154 mother-child pairs delivered at MSHS hospitals, covering 68,893 mothers and 93,154 children.^11-13^ The newborns in this cohort were born from 2007 to 2019, ensuring that all newborns had at minimum one year of follow-up (see also Supplemental Materials). This study was approved by the Mount Sinai institutional review board (IRB): IRB-20-01771.

### Digital phenotyping algorithm for rare genetic disorders

The *PheIndex* (Phenotype Index) digital phenotyping algorithm was developed based on 13 criteria that may be present in children with a rare genetic disorder. These criteria are primarily based on healthcare utilization patterns such as hospital encounters, procedures, specialist visits, and laboratory test orders. Orders that were subsequently cancelled were not considered. Additional criteria that were included were diagnostic codes of developmental delay and metabolic disease, and death. Description of the criteria with the associated scores is listed in Table 1.

**Table 1:** Description and scoring for the 13 *PheIndex* criteria

| Description | Scoring |
| --- | --- |
| *Prolonged stay in the neonatal intensive care unit (NICU) for term babies.* Full-term newborns who were admitted to the NICU and stayed for $\geq 4$ days. | Major; score = 3 |
| *Prolonged or multiple hospitalizations after discharged from birth.* Hospitalization is defined as inpatient stays with a duration $\geq 48$ hours. We included hospital stays where the calculated gestational age is older than 35 weeks and exclude the first newborn encounter if earlier than 35-week gestation. To meet this criterion, the patient must have either at least one prolonged hospitalization ($\geq 14$ days) or at least two hospitalizations ($\geq 48$ hours duration) for full-term or $\geq 3$ hospitalizations ($\geq 48$ hours) for pre-term babies. | Major; score = 3 |
| *Visits or consults with multiple specialists other than general pediatricians.* Twenty types of specialists were considered: Medical Genetics, Neurosurgery, Pediatric Allergy and Immunology, Pediatric Cardiology, Pediatric Dermatology, Pediatric Endocrinology, Pediatric GI/Pediatric Liver, Pediatric Hematology/Oncology, Pediatric Nephrology, Pediatric Neurology, Pediatric Ophthalmology, Pediatric Orthopedics, Pediatric Otolaryngology, Pediatric Pulmonology, Pediatric Rheumatology, Pediatric Surgery, Pediatric Urology, Transplant, Plastic Surgery. We counted the types of specialists each patient visited or consulted with and not the number of individual specialist visits. Preterm babies with $\geq 4$ types of specialists or full-term babies with $\geq 3$ types of specialists meet this criterion. We excluded Pediatric Infectious Disease specialty visits as infections in general are primarily due to environmental and non-genetic etiologies, and our aim was to identify a patient population enriched for children with genetic disorders. | Minor; score = 1 |
| *Multiple emergency room (ER) visits.* Full-term babies with $\geq 5$ ER visits **or** preterm babies with $\geq 7$ ER visits meet this criterion. | Minor; score = 1 |
| *Feeding support (Gastrostomy tube).* Patients who required feeding support were identified using ICD codes (Supplemental Table 2A) and procedures with description of “nasogastric”, “gastrostomy” and “feed”, or “gastrostomy” and “tube” in the procedure name. | Minor; score = 2 |
| *Respiratory support (tracheostomy and mechanical ventilation outside of surgery).* We used tracheostomy and ventilation (including CPAP) identified by procedure codes and diagnosis codes. If a surgical procedure was performed, the ventilatory support was required to be performed either 1 day before or 5 days after surgeries to be able to meet this criterion. | Minor; score = 2 |
| *Imaging.* We included patients that received computed tomography (CT) or magnetic resonance imaging (MRI) with completed encounter order status or preliminary/final results available from radiological exams. | Minor; score = 1 |
| *Genetic diagnostic tests.* We included patients who received genetic diagnostic tests such as gene sequencing or array comparative genomic hybridization regardless of test results. The records of genetic diagnostic tests were retrieved from procedure codes and labs. | Minor; score = 1 |
| *Metabolic diagnostic tests.* We included patients who received metabolic tests such as plasma amino acids panel or urine organic acids panel, regardless of test results. The records of metabolic diagnostic tests were retrieved from procedure codes and labs. | Minor; score = 1 |
| *In-hospital death.* Death information was retrieved from discharge location/disposition (expired, to funeral home/morgue or organ harvest) from encounter records. | Major; score = 3 |
| *Developmental delay.* Patients with developmental delay were identified by either a specialist visit with a developmental pediatrician or at least two occurrences of related ICD codes (Supplemental Table 2B) | Minor; score = 1 |
| *Diagnosis codes corresponding to metabolic diseases with $\geq 2$ encounters, (major, score=3).* We included patients with ICD codes for metabolic diseases (Supplemental Table 2C). | Major; score = 3 |
| *Heart surgeries, (major, score=3).*<br>Newborns that received heart surgeries were identified by encounters related to cardiothoracic surgeries or cardiothoracic intensive care unit (CTICU). | Major; score = 3 |

*PheIndex* combines these criteria in two different ways: (1) “*PheIndex Score*”, a quantified score indicating the severity of illness with a possible range between 0 and 24 generated by the sum of the score(s) associated with the criteria met by a child; and (2) “*PheIndex Classification*”, a binary classification of those who present with illness with an increased risk for genetic disorders (*PheIndex Classification* positive) if the following conditions are met: (a) ≥2 major criteria, (b) ≥1 major criteria and ≥1 minor criteria, (c) ≥5 minor criteria, or (d) deceased patient; or those who do not present illness with increased risk for genetic disorders (*PheIndex Classification* negative).

### Chart review verification of the PheIndex digital phenotyping

For the blinded chart review, we selected 200 charts consisting of children who were *PheIndex Classification* positive (N=100) and *PheIndex Classification* negative (N=100). We ensured that the 100 children who were negative covered quantified scores from 0 to 6 (inclusive), and from 3 to 21 for 100 children who were positive, based on the distribution of the *PheIndex Score*. Available records for this review were from encounters dated 01/01/2005 to 06/30/2020. All criteria determinations were based on available medical records up until three years of age. The review by the pediatrician had two steps: 1) validate the accuracy of the values assigned to each of the 13 criteria for each patient; and 2) summarize diagnostic information from the patient charts. The pediatrician had access to additional delivery notes, progress notes, admission/discharge summaries, and imaging notes. Information on diagnoses available in the notes documented by the pediatrician was then used by a clinical geneticist to decide whether the child presented with illness with an increased risk for genetic disorders. The possible categories of determination were: 1) “Definitively/possibly has genetic disorder diagnosis”, 2) “Does not have a genetic disorder”, 3) “Unknown, insufficient information to make determination on whether a genetic disorder was related with illness.”

### Statistical analysis

Full details are described in Supplemental Materials.

## Results

### Distribution of the 13 criteria in PheIndex

Our cohort included 93,154 newborns linked to 68,893 mothers who delivered in the MSHS from 2007 to 2019, with clinical features collected to 2020 (Table 2). We first assessed the frequency of each of the 13 *PheIndex* digital phenotyping criteria in our cohort and summarized the number of children aged 0 to 3 years old that satisfied each of the 13 criteria (Table 3). The most common criteria were multiple ER visits (3,919; 4.22%), followed by developmental delay (3,159; 3.39%), and multiple visits to specialists (3,091; 3.32%). The least common criteria were metabolic disease diagnosis codes (82; 0.09%) and feeding support (132; 0.14%). Figure 1A and 1B demonstrate the expected temporal relationship for achieving each criterion.

**Table 2:**
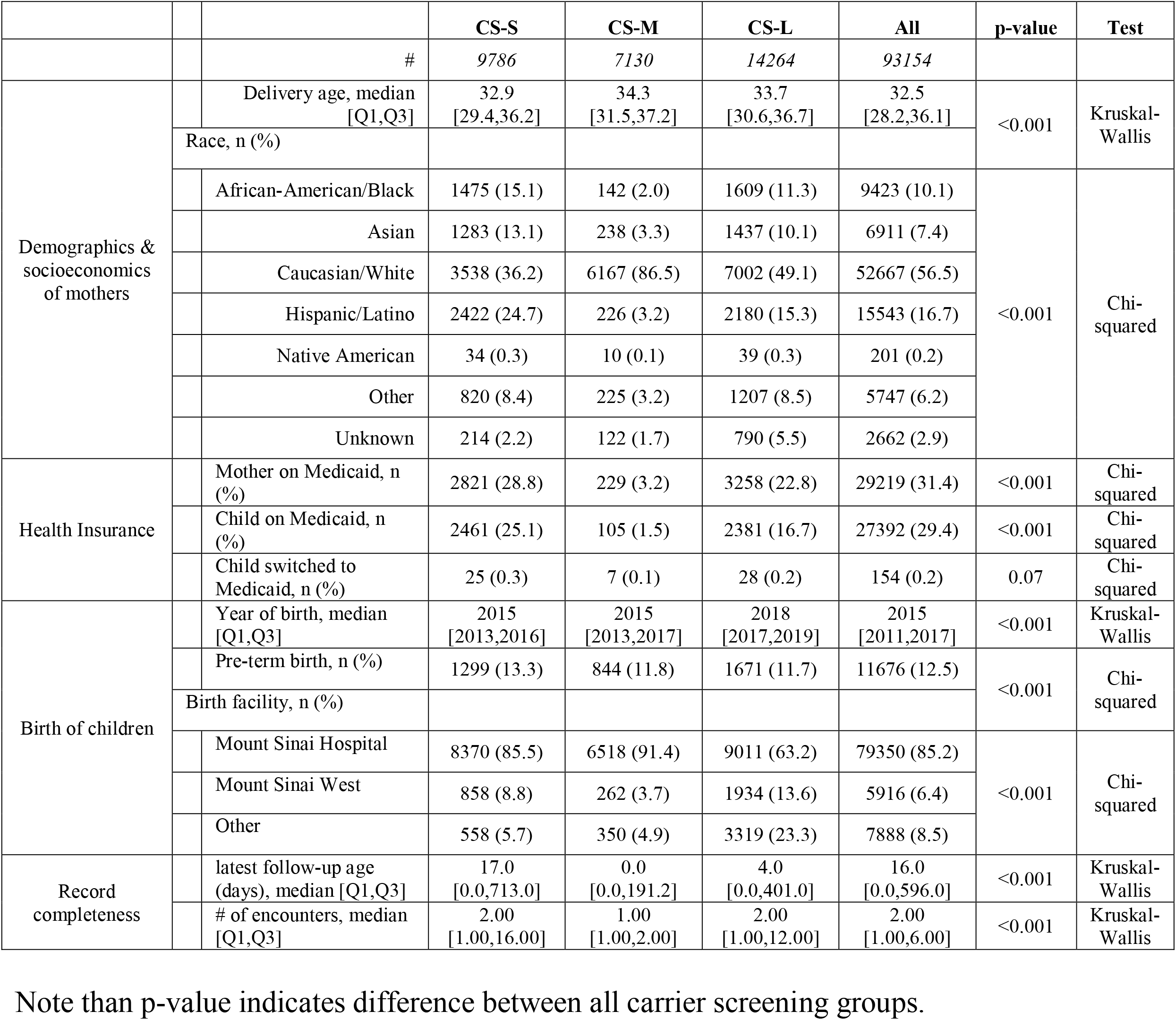
Demographic information of cohorts by carrier screening status.

|  |  | CS-S | CS-M | CS-L | All | p-value | Test |
| --- | --- | --- | --- | --- | --- | --- | --- |
|  | # | 9786 | 7130 | 14264 | 93154 |  |  |
| Demographics & socioeconomics of mothers | Delivery age, median [Q1,Q3] | 32.9 [29.4,36.2] | 34.3 [31.5,37.2] | 33.7 [30.6,36.7] | 32.5 [28.2,36.1] | <0.001 | Kruskal-Wallis |
|  | Race, n (%) |  |  |  |  |  |  |
|  | African-American/Black | 1475 (15.1) | 142 (2.0) | 1609 (11.3) | 9423 (10.1) | <0.001 | Chi-squared |
|  | Asian | 1283 (13.1) | 238 (3.3) | 1437 (10.1) | 6911 (7.4) |  |  |
|  | Caucasian/White | 3538 (36.2) | 6167 (86.5) | 7002 (49.1) | 52667 (56.5) |  |  |
|  | Hispanic/Latino | 2422 (24.7) | 226 (3.2) | 2180 (15.3) | 15543 (16.7) |  |  |
|  | Native American | 34 (0.3) | 10 (0.1) | 39 (0.3) | 201 (0.2) |  |  |
|  | Other | 820 (8.4) | 225 (3.2) | 1207 (8.5) | 5747 (6.2) |  |  |
|  | Unknown | 214 (2.2) | 122 (1.7) | 790 (5.5) | 2662 (2.9) |  |  |
| Health Insurance | Mother on Medicaid, n (%) | 2821 (28.8) | 229 (3.2) | 3258 (22.8) | 29219 (31.4) | <0.001 | Chi-squared |
|  | Child on Medicaid, n (%) | 2461 (25.1) | 105 (1.5) | 2381 (16.7) | 27392 (29.4) | <0.001 | Chi-squared |
|  | Child switched to Medicaid, n (%) | 25 (0.3) | 7 (0.1) | 28 (0.2) | 154 (0.2) | 0.07 | Chi-squared |
| Birth of children | Year of birth, median [Q1,Q3] | 2015 [2013,2016] | 2015 [2013,2017] | 2018 [2017,2019] | 2015 [2011,2017] | <0.001 | Kruskal-Wallis |
|  | Pre-term birth, n (%) | 1299 (13.3) | 844 (11.8) | 1671 (11.7) | 11676 (12.5) | <0.001 | Chi-squared |
|  | Birth facility, n (%) |  |  |  |  |  |  |
|  | Mount Sinai Hospital | 8370 (85.5) | 6518 (91.4) | 9011 (63.2) | 79350 (85.2) | <0.001 | Chi-squared |
|  | Mount Sinai West | 858 (8.8) | 262 (3.7) | 1934 (13.6) | 5916 (6.4) |  |  |
|  | Other | 558 (5.7) | 350 (4.9) | 3319 (23.3) | 7888 (8.5) |  |  |
| Record completeness | latest follow-up age (days), median [Q1,Q3] | 17.0 [0.0,713.0] | 0.0 [0.0,191.2] | 4.0 [0.0,401.0] | 16.0 [0.0,596.0] | <0.001 | Kruskal-Wallis |
|  | # of encounters, median [Q1,Q3] | 2.00 [1.00,16.00] | 1.00 [1.00,2.00] | 2.00 [1.00,12.00] | 2.00 [1.00,6.00] | <0.001 | Kruskal-Wallis |

Note than p-value indicates difference between all carrier screening groups.

**Table 3:** Number of children passing each individual *PheIndex* criteria.

| **PhenoIndex Criteria** | **n (%)** |
| --- | --- |
| multiple ER visits | 3919 (4.2) |
| developmental delay | 3159 (3.4) |
| multiple specialists | 3091 (3.3) |
| respiratory support | 2838 (3.0) |
| imaging | 1113 (1.2) |
| genetic tests | 704 (0.8) |
| prolonged in-patient stays | 500 (0.5) |
| metabolic tests | 448 (0.5) |
| death | 371 (0.4) |
| heart surgeries | 304 (0.3) |
| prolonged NICU stay | 279 (0.3) |
| feeding support | 132 (0.1) |
| metabolic ICD codes | 82 (0.1) |

**Figure 1.**
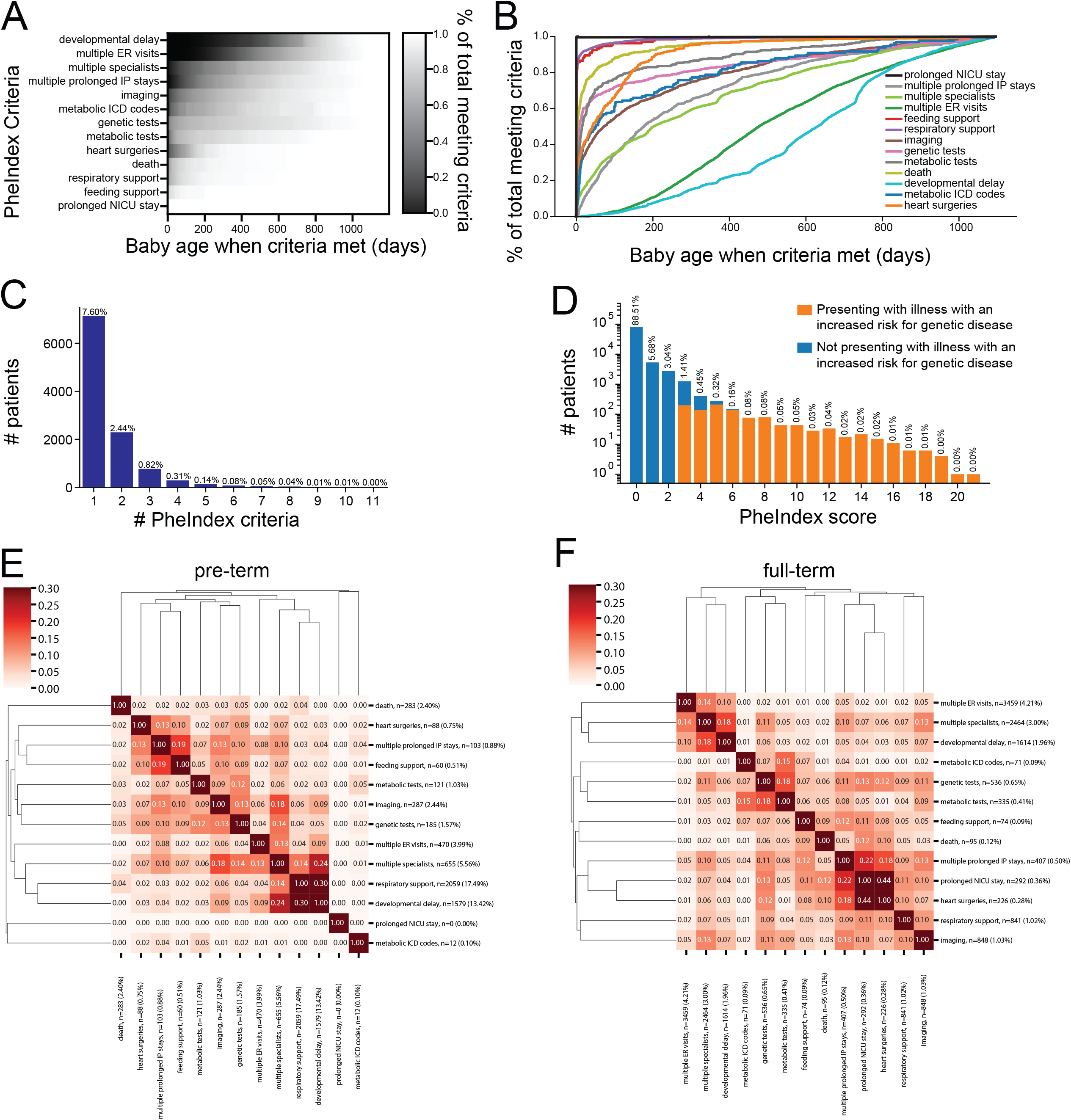
Distribution of *PheIndex* critera of children in the cohort. (A,B) Cumulative distribution of time when patients first meet each of the 13 *PheIndex* criteria. Only patients that met each criterion within the three-year limit were included in each cumulative distribution. (A) is sorted by the percentage of patients meeting the criteria at 200 days (least number of patients at the top). (C) Bar graph showing the showing the number and percentage of patients with passing different numbers of *PheIndex* criteria. (D) Distribution of *PheIndex* scores for children within the mother-child cohort. (E,F) Clustered heatmap showing the Jaccard index between possible pairs of *PheIndex* criteria in the pre-term (E) and full-term (F) cohorts. The number and percentage of patients for each criterion are labeled.

We generated a heatmap to show the number and percentage of patients who fell into different major and minor criteria combinations (Figure S1). The distribution for the total number of criteria for each child is given in Figure 1C. A large majority of patients (88.51%) did not meet any of the 13 criteria, and 98.55% met ≤2 criteria. We showed the distribution of *PheIndex Classification* – children who presented with illness with an increased risk for genetic disorders or not – stratified by the *PheIndex Score* (Figure 1D), as the *PheIndex Classification* depends on the specific combination of major and minor criteria for each patient. The majority of patients had a *PheIndex Score* ≤ 2 (97.23%), indicating that most children in our study population were not likely to have a rare genetic disorder. With our 13 criteria, the *PheIndex Classification* identified 1,088 children who were presenting with illness with an increased risk for genetic disorders out of 93,154 children (1.2%).

Hospital utilization patterns are known to vary between pre-term and full-term infants, e.g. pre-term infants often have more prolonged NICU stays. To assess this, we computed the similarity between all pairs of *PheIndex* criteria using the Jaccard index for each group (Figure 1E and 1F, Supplemental Materials). In the full-term cohort, *heart surgeries* and *prolonged NICU stay* had the highest Jaccard similarity of 0.44, in line with what we would expect to observe clinically. In the preterm cohort, prolonged NICU stay was not chosen to be a criterion because the majority of preterm infants have an extended NICU stay regardless of whether they have a rare genetic disorder or not.

### Validation of PheIndex: 13 Criteria and Overall Classification

First, we evaluated the accuracy of the values that were extracted from the EMR and assigned to the 13 different criteria for each patient, by comparing *PheIndex*’s identification of each of the 13 criteria against a pediatrician’s evaluation directly from the clinical notes for each patient, for a sample of 200 children (Table 4). The 200 children were sampled from those classified as presenting with illness with an increased risk for genetic disorders positive for a rare genetic condition (N=100) and those classified as negative (N=100). From this comparison, our digital phenotyping algorithm achieved an average accuracy of 94% across the 13 criteria. Accuracies were ≥90% for all criteria except for “*prolonged NICU stays”*, which yielded an accuracy of 81%.

**Table 4:** Accuracy of digital phenotype algorithm compared to chart review for individual *PheIndex* criteria.

| **PhenoIndex Criteria** | **Accuracy** |
| --- | --- |
| prolonged NICU stay | 81% |
| prolonged in-patient stays | 98% |
| multiple ER visits | 94% |
| multiple specialists | 93% |
| feeding support | 96% |
| respiratory support | 90% |
| imaging | 97% |
| genetic tests | 96% |
| metabolic tests | 96% |
| death | 98% |
| metabolic ICD codes | 97% |
| developmental delay | 93% |
| heart surgeries | 97% |

Next, we compared the *PheIndex Classification* against the classifications made by a pediatrician/medical geneticist (Table 5). Among the 200 children reviewed, 12 patients did not have sufficient clinical information for the medical geneticist to assess whether a genetic disorder may be present. Ten of these 12 patients were born extremely prematurely (born before 28 gestational weeks), which led to uncertainties as to whether the criteria that were met was because of prematurity or because of an underlying genetic disorder (as determined by the medical geneticist). Therefore, these 12 patients were excluded from this performance evaluation. Among the 188 patients remaining (88 classified as positive by *PheIndex* and 100 classified as negative), 85 patients were deemed to be true positives (definitively or possibly has a rare genetic disorder by chart review, 90% sensitivity/recall) and 91 patients were deemed to be true negatives (does not have a genetic disorder, 97% specificity). Three patients who were classified as positive by *PheIndex* were not thought to have a genetic disorder (false positive), and 9 patients were thought to definitively or possibly have genetic disorders but were classified as having no genetic disorders by *PheIndex* (false negative), yielding a positive predictive value (PPV) of 97%, negative predictive value (NPV) of 91%, and 94% accuracy. If we considered the prevalence of rare genetic disorders to be 3-3.6% of all livebirths,^14^ the adjusted PPV ranges from 48.1% to 48.3%.^15^

**Table 5:** Performance of *PheIndex* Classification against chart review.

| PhenoIndex<br>Classification |  | Clinical geneticist classification |  |  | Total |
| --- | --- | --- | --- | --- | --- |
|  |  | Does not have genetic disease | Definitively or possibly has genetic disease | Unknown (insufficient information) |  |
|  | Negative | 91 (True Negative) | 9 (False Negative) | 0 | 100 |
|  | Positive | 3 (False positive) | 85 (True Positive) | 12 | 100 |
|  | Total | 94 | 94 | 12 | 200 |

### PheIndex Scores by Carrier Screening Gene Panel Size

We examined the association between the *PheIndex Score*, an indicator of disease severity, and the three panel sizes (CS-S, CS-M, and CS-L). We first identified that 3 *PheIndex* criteria (multiple inpatient hospital stays, genetic testing, and developmental delay) were enriched for infants whose mothers had performed only CS-S testing compared with CS-M and CS-L (Table 6). For patients with at least 1 year of follow-up, we observed that the overall *PheIndex Scores* were higher in CS-S (mean=0.70) compared to CS-M (mean=0.38, p<0.001) and CS-L (mean=0.57, p<0.001) (Figure 3A); and CS-S (mean=0.85) compared to CS-M (mean=0.47, p<0.001, Student’s T-test) and CS-L (mean=0.70, p<0.001) for patients with at least 2 years of follow-up (Figure 3D).

**Table 6:** Number of children passing each individual *PheIndex* criteria split by carrier screen testing status.

|  | **CS-S** | **CS-M** | **CS-L** | **All** | **P-Value** | **Test** |
| --- | --- | --- | --- | --- | --- | --- |
| # | 9786 | 7130 | 14264 | 93154 |  |  |
| multiple ER visits, n (%) | 623 (6.4) | 25 (0.4) | 424 (3.0) | 3919 (4.2) | <0.001 | Chi-squared |
| developmental delay, n (%) | 515 (5.3) | 138 (1.9) | 522 (3.7) | 3159 (3.4) | <0.001 | Chi-squared |
| multiple specialists, n (%) | 497 (5.1) | 121 (1.7) | 431 (3.0) | 3091 (3.3) | <0.001 | Chi-squared |
| respiratory support, n (%) | 348 (3.6) | 201 (2.8) | 665 (4.7) | 2838 (3.0) | <0.001 | Chi-squared |
| imaging, n (%) | 132 (1.3) | 43 (0.6) | 162 (1.1) | 1113 (1.2) | <0.001 | Chi-squared |
| genetic tests, n (%) | 75 (0.8) | 31 (0.4) | 95 (0.7) | 704 (0.8) | 0.03 | Chi-squared |
| prolonged in-patient stays, n (%) | 51 (0.5) | 15 (0.2) | 45 (0.3) | 500 (0.5) | 0.002 | Chi-squared |
| metabolic tests, n (%) | 49 (0.5) | 26 (0.4) | 69 (0.5) | 448 (0.5) | 0.38 | Chi-squared |
| death, n (%) | 44 (0.4) | 10 (0.1) | 41 (0.3) | 371 (0.4) | 0.001 | Chi-squared |
| heart surgeries, n (%) | 23 (0.2) | 7 (0.1) | 17 (0.1) | 304 (0.3) | 0.03 | Chi-squared |
| prolonged NICU stay, n (%) | 24 (0.2) | 9 (0.1) | 27 (0.2) | 279 (0.3) | 0.22 | Chi-squared |
| feeding support, n (%) | 13 (0.1) | 3 (0.0) | 13 (0.1) | 132 (0.1) | 0.16 | Chi-squared |
| metabolic ICD codes, n (%) | 3 (0.0) | 4 (0.1) | 11 (0.1) | 82 (0.1) | 0.38 | Chi-squared |
| # of PheIndex criteria, mean (SD) | 0.24 (0.69) | 0.09 (0.41) | 0.18 (0.59) | 0.18 (0.63) | <0.001 | One-way ANOVA |
| PheIndex Score, mean (SD) | 0.31 (0.98) | 0.13 (0.62) | 0.24 (0.87) | 0.25 (0.97) | <0.001 | One-way ANOVA |

**Figure 2.**
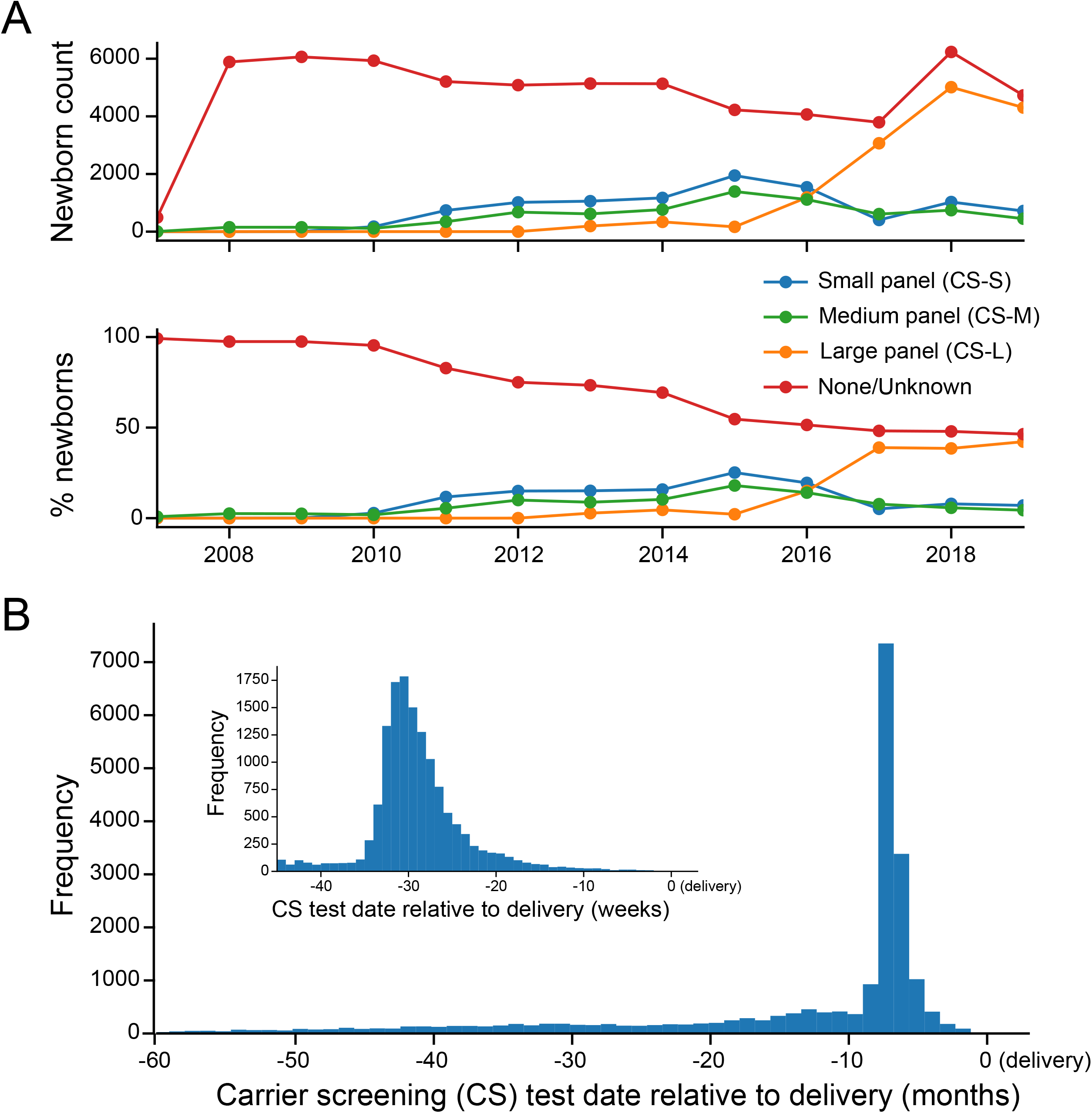
Summary statistics of the genetic carrier screening status for newborns in the mother-child cohort. (A) Top: Numbers of newborns whose mothers were tested with different genetic carrier screens arranged by years of birth (YOB). Bottom: Percentages of newborns whose mothers were tested with different genetic carrier screens arranged by YOB. (B) Histogram showing the distribution of genetic carrier tests dates (by month) relative to delivery (inset, weekly).

**Figure 3.**
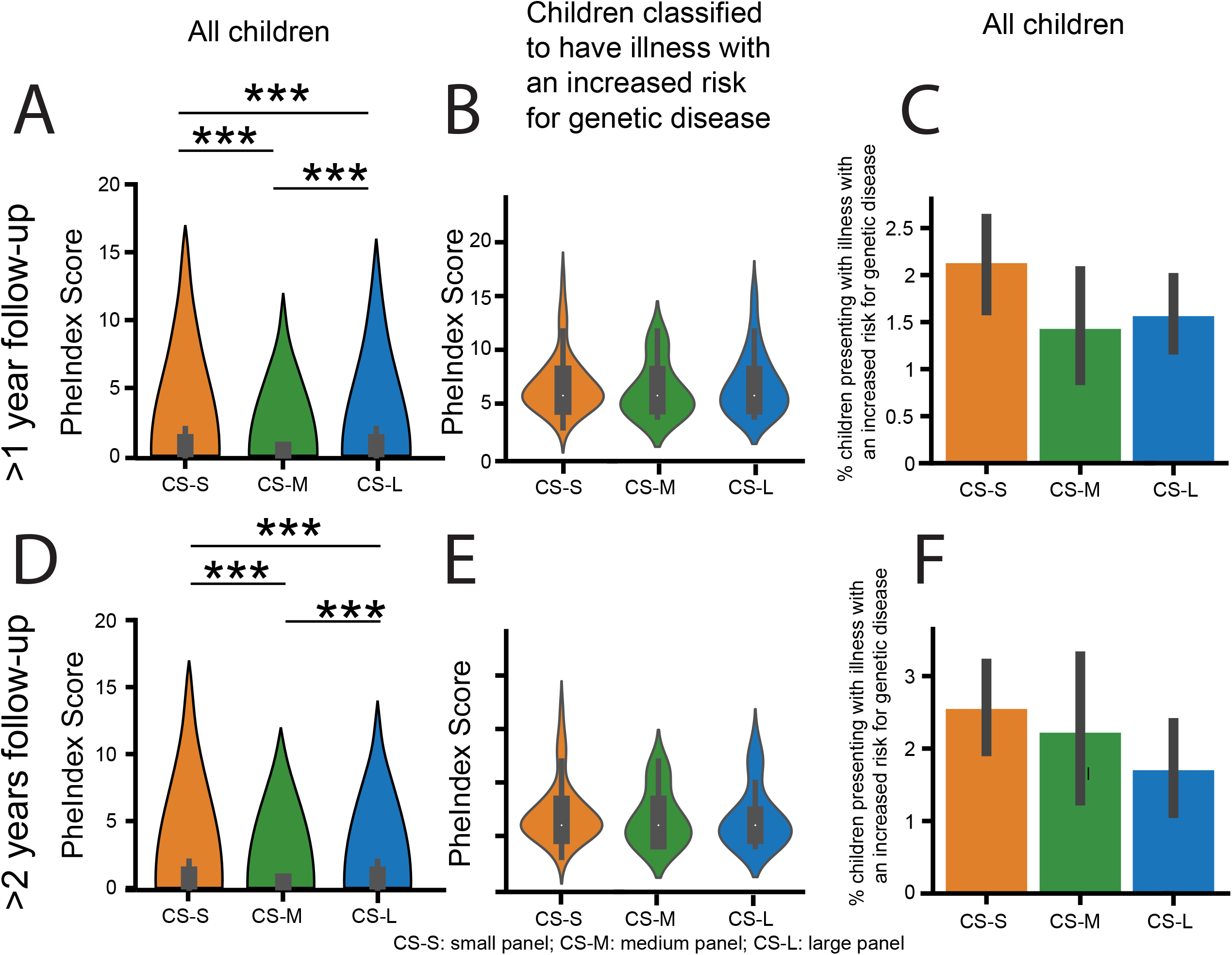
*PheIndex* Scores across the carrier screening (CS) testing cohort stratified by length of follow-up. (A) Average *PheIndex* score from the digital phenotype for all children with ≥1 year of follow-up in each CS testing cohort. Error bars show 95% confidence interval. *** denotes p<0.001. (B) Violin plot of *PheIndex* scores using the same categories as (A), but only including children labelled as “positive” from digital phenotype. (C) Percentage of children labelled as “positive” for each CS testing cohort. Error bars show 95% confidence interval. (D-F) Same as (A-C) but only including children with at least two years of follow-up.

### Comparison of time to onset for each criterion for CS-S and CS-L

To investigate the contributions of clinical factors to the *PheIndex* criteria and scores over time, we performed a sub-analysis between the CS-S and CS-L groups. We computed Kaplan-Meier survival curves for each of the 13 criteria for the CS-S and CS-L groups (Figure 4, Supplemental Figure 2) to examine the association of outcomes with time. We found that children in the CS-L cohort were less likely to see multiple specialists (p<0.001, Figure 4A), have multiple visits to the ER (p<0.001, Figure 4B), and less likely to undergo heart surgeries (p=0.06) or die early in childhood (p=0.058), compared to children in the CS-S cohort.

**Figure 4.**
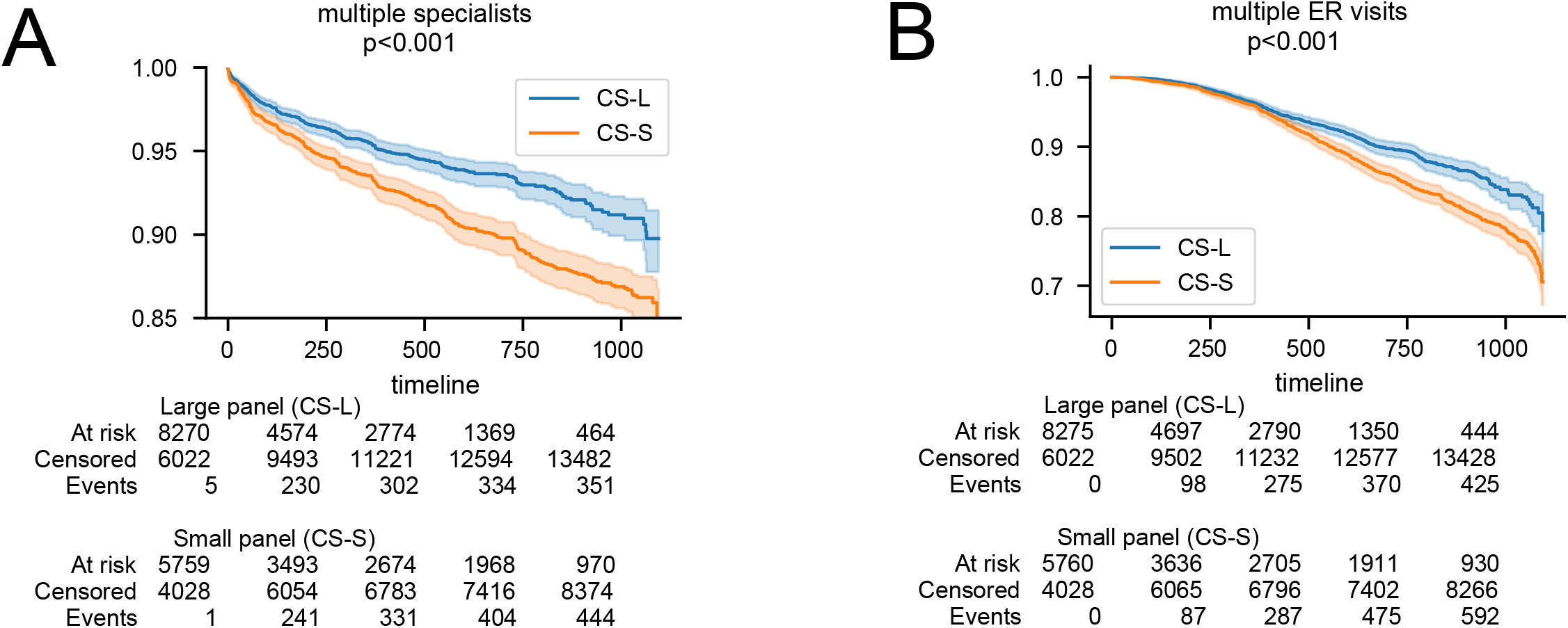
Kaplan-Meier curves stratified by carrier screening panel size, Carrier screening, small panel / CS-S (blue) vs Carrier screening, large panel / CS-L (orange). (A) Shows that children whose mothers received CS-S met the multiple specialist criterion in a greater proportion than those whose mothers received CS-L at three years of follow-up. (B) Shows that children whose mothers received CS-S met the multiple ER visits criterion in a greater proportion than those whose mothers received CS-L at three years of follow-up. Shaded areas denote 95% confidence interval.

### Regression Analysis

In the Cox proportional hazards model, we found that for children whose mothers were administered a CS-L panel, a 36% (p=0.005) reduction of being classified as presenting with illness with an increased risk for genetic disorders was estimated, compared to the children whose mothers ordered were administered a CS-S panel test (Supplemental Figure 4).

## Discussion

Identifying pediatric patients across an entire population with or who possibly has a rare genetic disorder is critical for improving patient outcomes. We and others have attempted to identify patients with specific genetic disorders using EMR data, but have found that such a process is not straightforward, largely due to coding differences, unconfirmed diagnoses, variation in disease names and terminology, and inaccurate information represented in medical records.^16,17^ For most rare genetic disorders, it is difficult to identify patients with specific genetic disorders, given ICD codes are often nonspecific.^1,18,19^ Additionally, seeking to analyze individual diseases, even in EMR databases with millions of patients, would result in underpowered studies given the low frequency of individual rare genetic disorders. However, by using a global metric as opposed to ones derived from specific individual diseases, we were able to identify a large cohort that provided for sufficient statistical power to assess the association of differently sized CS panels with risk of genetic disorders.

In this study, we developed a novel, rule-based digital phenotyping algorithm (*PheIndex*) that utilizes 13 criteria to derive a *PheIndex Score* for children from birth to 3 years of age, in order to classify whether a child is presenting with an illness that may be a rare genetic disorder. Importantly, our score is an evaluation of overall health rather than the presence of specific features of individual diseases. To our knowledge, such an approach has not been developed previously. The criteria for the *PheIndex Score* include items that could be extracted from the EMR with a high degree of precision and accuracy. *Our PheIndex Score* may be utilized for various purposes, including its use as a clinical guide to shorten the diagnostic odyssey of hard-to-diagnose patients, timely administration of therapeutics by facilitating more rapid diagnosis, and/or assessing clinical benefit of genetic testing, all of which help enable the practice of precision medicine in a way that may be more accessible to all. Chart review from clinical genetics experts, confirmed that our *PheIndex* algorithm has the following performance characteristics when the numbers of cases and controls are equal: precision of 97%, recall of 90%, and accuracy of 94%.

To demonstrate the ability of our algorithm to identify an enriched set of patients at risk of harboring a rare genetic condition, we leveraged carrier screening results in mothers who delivered a baby in a large health system. We examined the association between a mother’s carrier screening panel size and *PheIndex Score*. We found that CS-L was not only associated with a lower overall *PheIndex Score*, but was also significantly associated with a decreased need for a child having multiple specialist visits and multiple ER visits. In our sub-analysis using a cohort of mother-child pairs whose mother received CS-L or CS-S and with whom the child received at least two years of follow-up, we noted that those in the CS-L group reached the criteria of *multiple specialists* and *genetic diagnostic tests* earlier than those in the CS-S group. This result is notable as it supports that administration of a CS-L panel test may enable earlier diagnosis of genetic disorders in children. Alternatively, testing using a CS-L panel may increase awareness of parental carrier status, thus enabling prenatal diagnostic testing for a larger number of conditions. This increased awareness may also lead to early referral of children manifesting severe illness for rare genetic diagnostic testing and subsequent referrals to the appropriate specialists and potentially earlier treatment. Parental carrier status may also lead to earlier postnatal diagnostic genetic testing and thus confirmation of a particular genetic disorder.

### Limitations

While our study population is likely representative of other large, diverse metropolitan areas, it may be less representative of smaller-sized cities and rural areas. Also, we provided an adjusted PPV of 48% based on an estimated prevalence of rare genetic disorders in the general population. However, precise estimates of rare genetic disorder prevalence are unavailable, and may also not reflect the PPV for the target population of our algorithm (i.e. children aged 0-3) due to differences in age of onset.^14^ Another potential limitation of our study is that we used only de-identified data available in structured EMR databases, and thus did not include all the information that would be available to physicians, such as clinical notes. However, despite not having access to all available clinical notes, our digital phenotype agreed with physician chart review 94% of the time (under conditions in which the number of cases and controls were sampled to be the same), proving that our algorithm successfully identifies children possibly with rare genetic disorders. In the few occasions where there were discrepancies, this was typically due to incomplete documentation of orders, such as respiratory support and feeding support in *PheIndex* negative children that was uncovered in the notes during chart review. Thus, we believe that our digital phenotype will be more accurate using the *PheIndex* criteria extracted from notes in addition to structured EMR data. With respect to demonstrating the application of *PheIndex* across groups receiving comprehensive carrier screening and those receiving reduced or no carrier screening, the MSHS is unique in that beginning in 2016, CS-L was offered to all women considering pregnancy or already pregnant regardless of the mother’s ability to pay or health insurance coverage. Notably, our cohort is comprised of linked mother-child pairs and thus does not directly assess the rate of mothers who chose not to continue pregnancies with an affected fetus; however, the improvement in health of children whose mothers received CS-L may be due to couples choosing to proceed with various reproductive health options such as *in vitro* fertilization (IVF), in order to reduce the chances of having a child affected with one of as many as 283 genetic conditions assessed in the CS-L panel described in our study. Additionally, mothers who chose to receive CS-L may be more likely to complete additional genetic testing via chorionic villus sampling or amniocentesis in the setting of advanced maternal age or family history of genetic disorders. Moreover, while we controlled for differential follow-up time and likely confounders, there may still be unmeasured confounding in the Cox regression model.

## Conclusions

In summary, we utilized a comprehensive EMR to develop a novel digital phenotyping algorithm for identification of a pediatric population with a definitive or possible genetic disorder. Our method utilizes a global approach as opposed to identifying patients in the EMR with each specific genetic disorder, which is fraught for misdiagnoses and error. In addition, our study is the first with adequate sample size and follow up to evaluate the health of children from birth to 3 years of age. Using a mother-child cohort that links children to mothers’ genetic carrier screening status, we have identified that *PheIndex Scores* are lower at one or two years of follow-up in children whose mothers received CS-L relative to CS-S. We believe that our *PheIndex* algorithm will address an unmet need to identify children with rare genetic disorders and potentially help overcome well-known obstacles such as underdiagnosis and delayed diagnosis.^20^

## Data Availability

The clinical data used in this study is under license from Mount Sinai Data Warehouse. As a result, this dataset is not publicly available. Qualified researchers affiliated with the Mount Sinai Health Systems may apply for access to these data through the Mount Sinai Health Systems Institutional Review Board.

## Abbreviations

CSER: Clinical Sequencing Exploratory Research
CS: carrier screening
CS-L: carrier screening, large panel
CS-M: carrier screening, medium panel
CS-S: carrier screening, small panel
CT: computed tomography
CTICU: cardiothoracic intensive care unit
eMERGE: Electronic Medical Records & Genomics EHR
EMR: electronic medical record
ER: emergency room
ICD: International Classification of Diseases
MRI: magnetic resonance imaging
MSHS: Mount Sinai Health System
NICU: neonatal intensive care unit
NPV: negative predictive value
PPV: positive predictive value

## ACKNOWLEDGEMENTS

We would like to thank Drs. Mitchell K. Higashi and Paul Kruszka for reviewing the manuscript and Mount Sinai Data Warehouse for EMR data. We also thank the GeneDx IT team for infrastructural and computational support.

## Notes

**Conflict of Interest Disclosures:** The authors have no conflicts of interest relevant to this article to disclose.

**Funding/Support:** None. This project was performed in collaboration with GeneDx. GeneDx is a company that integrates genetic testing and data analytics to improve diagnosis, treatment, and prevention of disease. The Icahn School of Medicine at Mount Sinai holds equity in this for profit company.

### Competing Interest Statement

BDW, LYL, RAS, DC, LS, SL, JT, SL, ZW, GS, LE, RC, EES, LL are formerly or currently employed by GeneDx. GeneDx is a company that integrates genetic testing and data analytics to improve diagnosis, treatment, and prevention of disease. The Icahn School of Medicine at Mount Sinai holds equity in this for-profit company.

### Funding Statement

This study did not receive any funding.

### Author Declarations

Ethics committee/IRB of Mount Sinai Health Systems gave approval for this work (IRB-20-01771).

