## Supplemental Materials for "An algorithm to identify patients with rare genetic disorders and its real-world data application"

**METHODS**

*Construction of mother-child cohort from the MSHS EMR*

To accurately ascertain the gestational week at birth and determine the term status of a newborn, a mother’s EMR had to be linked. In other words, we identified the mother-child pairs, where we obtained mothers’ delivery records for pregnant women who delivered in the MSHS and linked their corresponding newborn with the pregnancy and delivery journey. In total, we identified 93,154 mother-child pairs delivered at MSHS hospitals, covering 68,893 mothers and 93,154 children. Moreover, for the children, we obtained gestational age and all diagnoses, procedures, vital signs, laboratory tests, and medications available in the EMR from birth until any subsequent hospital encounters of any type up to three years of age. For demonstrating the application of the digital phenotyping algorithm, we extracted additional demographic information for the mothers including self-reported race and age at delivery as well as socioeconomic features such as Medicaid status.

*Digital phenotyping algorithm for rare genetic disorders*

The *PheIndex* (Phenotype Index) digital phenotyping algorithm was developed based on 13 criteria that may be present in children with a rare genetic disorder. These criteria are primarily based on healthcare utilization patterns such as hospital encounters, procedures, specialist visits, and laboratory test orders. Orders that were subsequently cancelled were not considered. Additional criteria that were included were diagnostic codes of feeding support, developmental delay, and metabolic disease (see Supplemental Table 1A-C), and death. The cut-off for each criterion was chosen based on the distribution in the population with reference to clinical relevance in identifying children with rare genetic disorders. Thus, the aim of this algorithm is to capture a patient population enriched with rare genetic disorders. Based on the severity of illness reflected by each rule, we classified 5 out of these 13 criteria as “major” and the remaining 8 as “minor”, as well as a score for each criterion scaled from 1 to 3, to account for the severity of illness in a clinical setting. A score of 3 indicates a criterion correlating with more severe illness, whereas a score of 1 reflects less severe illness. Four criteria take into account term status (pre-term or full-term).

*Chart review verification of the PheIndex digital phenotyping*

To assess the accuracy of our *PheIndex* digital phenotyping algorithm, manual chart review was conducted in a blinded fashion for the validation of the 13 criteria listed above. Since we used structured, deidentified data for developing the digital phenotyping algorithm, full clinical information may not be present, particularly for specific clinical features noted in free text format in clinical notes. Therefore, blinded chart review by physicians is necessary. In the chart review process, a pediatrician examined all the clinical data for each patient including medical history such as birth history; all encounters including corresponding notes for outpatient, emergency department, and inpatient care; lab orders; imaging studies; and death records within the hospital medical records system to ascertain the presence of clinical criteria that comprise our *PheIndex* digital phenotyping.

For the blinded chart review, we selected 200 charts consisting of children who were *PheIndex Classification* positive (N=100) and *PheIndex Classification* negative (N=100). Moreover, we ensured that the 100 children who were negative covered quantified scores from 0 to 6 (inclusive), and from 3 to 21 for 100 children who were positive, based on the distribution of the *PheIndex* quantified score (see Figure 1D in main text). Available records for this review were from encounters dated 01/01/2005 to 06/30/2020. All criteria determinations were based on available medical records up until three years of age. Chart selection covered each rule that we used to identify the phenotype to ensure representation including gestational age, NICU stay, emergency room visits, hospitalizations and duration of hospitalizations, subspecialty visits/consultations, presence of gastrostomy tube, presence of tracheostomy tube or utilization of mechanical ventilation in the absence of surgery, CT or MRI imaging studies, metabolic testing, genetic testing, metabolic disease diagnosis, developmental delay, prior cardiac surgery, and death.

An application of PheIndex in health services research with real-world data

*Utilization, extraction, and categorization of genetic carrier screening status for mothers*

We identified whether mothers received any genetic carrier screening (CS) tests and the type of test during pregnancy or prior to conception by mining Sema4’s laboratory’s test database that was built using the laboratory information system partner, Medgis (http://www.medgis.net/index.html). We manually categorized the types of CS tests into the following three groups: (1) CS Small panel (CS-S), which covers one to four diseases: *CFTR* (cystic fibrosis), *SMN1/SMN2* (spinal muscular atrophy), *FMR1* (fragile X syndrome) and *DHCR7* (Smith-Lemli-Opitz syndrome), (2) CS Medium panel (CS-M): which covers 5 to 99 genes including test panels designed for those with Jewish ancestry such as the Sema4 Ashkenazi Jewish Panel (36 diseases), and (3) CS Large panel, (CS-L): which covers ≥100 genes, including Sema4 Expanded Carrier Screening 283 genes panel, with a cut-off of 1 in 200 for any ethnicity for the carrier frequency for autosomal recessive diseases and with criteria established for disease severity and penetrance. A complete list of 283 genes with corresponding conditions is shown in Supplemental Table 2.

*Statistical analysis*

The Jaccard index was computed to examine the intersection of children between pairs of the *PheIndex* criteria: 13 for full-term and 12 for preterm newborns (prolonged NICU stay was not a criteria as preterm newborns stay in NICU for being preterm and not necessarily related to a rare disorder diagnosis). We described continuous variables as their median and quantile range, and categorical variables as a number and percentage. We performed statistical tests by ANOVA or two sample t-test for continuous variables and Chi-square test for categorical variables, respectively. Since the CS-L test was developed and offered to MSHS patients starting in 2016, there is a shorter follow-up window compared with those who were administered the CS-S test. Therefore, we also stratified *PheIndex Scores* by length of follow-up. To investigate the clinical factors that contribute to higher / lower scores in children over time, we performed a sub-analysis between the CS-S and CS-L subcohorts and computed Kaplan-Meier survival curves for each of the 13 criteria.

To assess the statistical association between CS status and *PheIndex Classification* and adjust for follow-up time, we fit a Cox proportional hazards model and controlled for preterm birth, newborns’ year of birth (YOB), number of well-child visits, whether a mother had Medicaid, and mothers’ self-reported racial groups.

We performed all analyses using R (version 3.6.1) and Python (version 3.7). We considered p<0.05 as statistically significant.

**RESULTS**

*Distribution of the 13 criteria in PheIndex by preterm status*

We note that hospital utilization patterns are known to vary between pre-term and full-term infants, since pre-term infants often have more clinical needs and prolonged NICU stays. To assess this, for each group we computed the similarity between all pairs of *PheIndex* criteria using the Jaccard index (Figure 1E and 1F in main text). In the full-term cohort, *heart surgeries* and *prolonged NICU stay* had the highest Jaccard similarity of 0.44, in line with what we would expect to observe clinically. In the preterm cohort, prolonged NICU stay was not chosen to be a criterion because the majority of preterm infants have an extended NICU stay regardless of whether they have a rare genetic disorder or not. Additionally, we note that *multiple specialists* and *developmental delay* had a Jaccard similarity coefficient of 0.24 (the second highest ranked similarity) in the pre-term cohort and 0.18 (the third highest ranked similarity) in the full-term cohort, consistent with standard clinical practice in which those with developmental delay are referred to specialists such as developmental pediatricians, pediatric neurologists, and/or clinical geneticists.

*Descriptive statistics to demonstrate real-world application of PheIndex*

In this cohort, 9786, 7130, and 14,264 mothers received carrier screening comprised of panels that included ≤4 genes (Small; CS-S), 5-99 genes (Medium; CS-M), and ≥100 genes (Large; CS-L), respectively. Mothers with self-reported races of African American/Black, Asian, or Hispanic/Latino, as well as families where mothers or children were on Medicaid, were significantly more likely to have a CS-S panel tests compared to CS-M or CS-L panel tests (p<0.001, Chi-squared test, Table 1 in main text). Further, CS-M panel tests were over-represented in mothers who self-identified as Caucasian/White, reflecting the historical origins of CS testing in the MSHS that had been focused on reproductive couples with Ashkenazi Jewish ancestry. Beyond race and socioeconomic status, we found that a mother’s delivery age, the average follow-up time, and the distribution of patients across the different health facilities in the MSHS, all varied significantly across the three CS panel size categories (Table 1 in main text).

*Temporal trends in CS testing*

Our cohort includes babies born between 2007 and 2019, and our clinical information was collected up to 2020 to ensure at least one year of follow-up on the children in the cohort. Over this time span, the uptake of CS-S, CS-M and CS-L panels varied as new technologies and our understanding of monogenic disorders dramatically increased. As a result, larger panels became available over time and were adopted into clinical practice. CS-S and CS-M testing grew steadily from 2010 to 2015 (Figure 2A) but were followed by the development of more comprehensive panels (CS-L) in 2015. CS-L was rapidly adopted as the standard of care in the MSHS, with CS-S or CS-M panel testing decreasing beyond 2015 while CS-L panel testing increased.

We also examined the timing of CS testing relative to the delivery date (Figure 2B). For the majority of patients (52.8%), CS was performed in the first trimester of pregnancy. For 36.0% of patients, CS testing was performed earlier than 10 months prior to delivery, and therefore was likely performed in a previous pregnancy or as part of pre-conception planning. The remaining 11.2% of patients had CS testing performed after their first trimester, but before delivery.

*Comparison of time to onset for each criterion for CS-S and CS-L*

In a sub-analysis of patients who had at least two years of follow-up, we assessed the age of onset for each of the 13 criteria within the follow up period and found that the time to meeting *multiple specialists* criteria was significantly less in the CS-L group (median = 148 [41, 339] days) than the CS-S group (median = 192 [55, 394]; p=0.048, Mann-Whitney U-test; Supplemental Figure 3A). Another criterion, ordering *genetic diagnostic tests,* also tended to occur earlier for children in the CS-L group (median = 66 [4, 72] days) compared to the CS-S group (median = 161 [2, 208] days; p=0.051, Mann-Whitney U-test; Supplemental Figure 3B).
