## Supplemental Tables for "An algorithm to identify patients with rare genetic disorders and its real-world data application"

**Supplemental Table 1A: Gastronomy ICD codes**

| V44.1 (ICD9) | Gastrostomy status |
| --- | --- |
| 779.31 (ICD9) | Feeding problems in newborn |
| Z93.1 (ICD10) | Gastrostomy status |
| P92.9 (ICD10) | Feeding problem of newborn, unspecified |

**Supplemental Table 1B: Developmental delay ICD codes**

| 315.3 (ICD9) | Expressive language disorder |
| --- | --- |
| 315.31 (ICD9) | Expressive language disorder |
| 315.32 (ICD9) | Mixed receptive-expressive language disorder |
| 315.39 (ICD9) | Other developmental speech or language disorder |
| 315.4 (ICD9) | Developmental coordination disorder |
| 315.8 (ICD9) | Other specified delays in development |
| 783.40 (ICD9) | Lack of normal physiological development |
| V40.0 (ICD9) | Mental and behavioral problems with learning |
| R62.50 (ICD10) | Developmental delay |
| F80.1 (ICD10) | Expressive language delay |
| F80.2 (ICD10) | Receptive expressive language disorder |
| F80.9 (ICD10) | Speech developmental delay |
| F81.9 (ICD10) | Cognitive developmental delay |
| F82 (ICD10) | Developmental delay of gross and fine motor function |
| F88 (ICD10) | Global developmental delay |
| Z87.898 (ICD10) | History of developmental delay |

**Supplemental Table 1C: Diagnosis codes corresponding to metabolic diseases**

| E70* (excl. E70.331: Hermansky-Pudlak syndrome) | Disorders of aromatic amino-acid metabolism |
| --- | --- |
| E71* | Disorders of branched-chain amino-acid metabolism and fatty-acid metabolism |
| E72* | Other disorders of amino-acid metabolism |
| E74* | Other disorders of carbohydrate metabolism |
| E75* | Disorders of sphingolipid metabolism and other lipid storage disorders |
| E76* | Disorders of glycosaminoglycan metabolism |
| E78* | Disorders of lipoprotein metabolism and other lipidemias |
| D81.810 | Biotinidase deficiency |

**Supplemental Table 2: 283 genes/conditions on the large carrier screening panel (CS-L)**

| **GeneSymbol** | **GeneID** | **Disease name** | **Inheritance** |
| --- | --- | --- | --- |
| ABCB11 | 8647 | progressive_familial_intrahepatic_cholestasis | AR |
| ABCC8 | 6833 | familial_hyperinsulinism | AR |
| ABCD1 | 215 | adrenoleukodystrophy | XL |
| ACAD9 | 28976 | mitochondrial_complex_i_deficiency | AR |
| ACADM | 34 | medium_chain_acyl-coa_dehydrogenase_deficiency | AR |
| ACADVL | 37 | very_long_chain_acyl-coa_dehydrogenase_deficiency | AR |
| ACAT1 | 38 | beta-ketothiolase_deficiency | AR |
| ACOX1 | 51 | acyl-coa_oxidase_i_deficiency | AR |
| ACSF3 | 197322 | combined_malonic_and_methylmalonic_aciduria | AR |
| ADA | 100 | adenosine_deaminase_deficiency | AR |
| ADAMTS2 | 9509 | ehlers-danlos_syndrome | AR |
| AGA | 175 | aspartylglycosaminuria | AR |
| AGL | 178 | glycogen_storage_disease | AR |
| AGPS | 8540 | rhizomelic_chondrodysplasia_punctata | AR |
| AGXT | 189 | primary_hyperoxaluria | AR |
| AIRE | 326 | autoimmune_polyendocrinopathy | AR |
| ALDH3A2 | 224 | sjogren-larsson_syndrome | AR |
| ALDOB | 229 | hereditary_fructose_intolerance | AR |
| ALG6 | 29929 | congenital_disorder_of_glycosylation | AR |
| ALMS1 | 7840 | alstrom_syndrome | AR |
| ALPL | 249 | hypophosphatasia | AR |
| AMT | 275 | glycine_encephalopathy | AR |
| AQP2 | 359 | nephrogenic_diabetes_insipidus | AR |
| ARSA | 410 | metachromatic_leukodystrophy | AR |
| ARSB | 411 | mucopolysaccharidosis | AR |
| ASL | 435 | argininosuccinic_aciduria | AR |
| ASNS | 440 | asparagine_synthetase_deficiency | AR |
| ASPA | 443 | canavan_disease | AR |
| ASS1 | 445 | citrullinemia | AR |
| ATM | 472 | ataxia-telangiectasia | AR |
| ATP6V1B1 | 525 | renal_tubular_acidosis | AR |
| ATP7A | 538 | menkes_disease | XL |
| ATP7B | 540 | wilson_disease | AR |
| ATRX | 546 | alpha-thalassemia_mental_retardation_syndrome | XL |
| BBS1 | 582 | bardet-biedl_syndrome | AR |
| BBS10 | 79738 | bardet-biedl_syndrome | AR |
| BBS12 | 166379 | bardet-biedl_syndrome | AR |
| BBS2 | 583 | bardet-biedl_syndrome | AR |
| BCKDHA | 593 | maple_syrup_urine_disease | AR |
| BCKDHB | 594 | maple_syrup_urine_disease | AR |
| BCS1L | 617 | gracile_syndrome | AR |
| BLM | 641 | bloom_syndrome | AR |
| BSND | 7809 | bartter_syndrome | AR |
| BTD | 686 | biotinidase_deficiency | AR |
| CAPN3 | 825 | limb-girdle_muscular_dystrophy | AR |
| CBS | 875 | homocystinuria | AR |
| CDH23 | 64072 | Usher syndrome, type ID | AR |
| CEP290 | 80184 | leber_congenital_amaurosis | AR |
| CERKL | 375298 | retinitis_pigmentosa | AR |
| CFTR | 1080 | cystic_fibrosis | AR |
| CHM | 1121 | choroideremia | XL |
| CHRNE | 1145 | congenital_myasthenic_syndrome | AR |
| CIITA | 4261 | bare_lymphocyte_syndrome,_type_ii | AR |
| CLN3 | 1201 | neuronal_ceroid-lipofuscinosis | AR |
| CLN5 | 1203 | neuronal_ceroid-lipofuscinosis | AR |
| CLN6 | 54982 | neuronal_ceroid-lipofuscinosis | AR |
| CLN8 | 2055 | neuronal_ceroid-lipofuscinosis | AR |
| CLRN1 | 7401 | usher_syndrome | AR |
| CNGB3 | 54714 | achromatopsia | AR |
| COL27A1 | 85301 | steel_syndrome | AR |
| COL4A3 | 1285 | alport_syndrome | AR |
| COL4A4 | 1286 | alport_syndrome | AR |
| COL4A5 | 1287 | alport_syndrome | XL |
| COL7A1 | 1294 | dystrophic_epidermolysis_bullosa | AR |
| CPS1 | 1373 | carbamoyl_phosphate_synthetase_i_deficiency | AR |
| CPT1A | 1374 | carnitine_palmitoyltransferase_ia_deficiency | AR |
| CPT2 | 1376 | carnitine_palmitoyltransferase_ii_deficiency | AR |
| CRB1 | 23418 | leber_congenital_amaurosis | AR |
| CTNS | 1497 | cystinosis | AR |
| CTSK | 1513 | pycnodysostosis | AR |
| CYBA | 1535 | chronic_granulomatous_disease | AR |
| CYBB | 1536 | chronic_granulomatous_disease | XL |
| CYP11B2 | 1585 | corticosterone_methyloxidase_type_i_deficiency | AR |
| CYP17A1 | 1586 | congenital_adrenal_hyperplasia | AR |
| CYP19A1 | 1588 | aromatase_deficiency | AR |
| CYP21A2 | 1589 | congenital_adrenal_hyperplasia | AR |
| CYP27A1 | 1593 | cerebrotendinous_xanthomatosis | AR |
| DCLRE1C | 64421 | omenn_syndrome | AR |
| DHCR7 | 1717 | smith-lemli-opitz_syndrome | AR |
| DHDDS | 79947 | retinitis_pigmentosa | AR |
| DLD | 1738 | pyruvate_dehydrogenase_deficiency | AR |
| DMD | 1756 | duchenne_and_becker_muscular_dystrophy | XL |
| DNAH5 | 1767 | primary_ciliary_dyskinesia | AR |
| DNAI1 | 27019 | primary_ciliary_dyskinesia | AR |
| DNAI2 | 64446 | primary_ciliary_dyskinesia | AR |
| DYSF | 8291 | limb-girdle_muscular_dystrophy | AR |
| EDA | 1896 | hypohidrotic_ectodermal_dysplasia | XL |
| EIF2B5 | 8893 | leukoencephalopathy_with_vanishing_white_matter | AR |
| EMD | 2010 | emery-dreifuss_muscular_dystrophy | XL |
| ESCO2 | 157570 | roberts_syndrome | AR |
| ETFA | 2108 | glutaric_acidemia | AR |
| ETFDH | 2110 | glutaric_acidemia | AR |
| ETHE1 | 23474 | ethylmalonic_encephalopathy | AR |
| EVC | 2121 | ellis-van_creveld_syndrome | AR |
| EYS | 346007 | retinitis_pigmentosa | AR |
| F11 | 2160 | congenital_factor_xi_deficiency | AR |
| F9 | 2158 | factor IX deficiency (X-linked) | XL |
| FAH | 2184 | tyrosinemia | AR |
| FAM161A | 84140 | retinitis_pigmentosa | AR |
| FANCA | 2175 | fanconi_anemia | AR |
| FANCC | 2176 | fanconi_anemia | AR |
| FANCG | 2189 | fanconi_anemia | AR |
| FH | 2271 | fumarase_deficiency | AR |
| FKRP | 79147 | limb-girdle_muscular_dystrophy | AR |
| FKTN | 2218 | familial_dilated_cardiomyopathy | AR |
| FMR1 | 2332 | fragile_x_syndrome | XL |
| G6PC | 2538 | glycogen_storage_disease | AR |
| GAA | 2548 | glycogen_storage_disease | AR |
| GALC | 2581 | krabbe_disease | AR |
| GALK1 | 2584 | galactokinase deficiency | AR |
| GALT | 2592 | galactosemia | AR |
| GAMT | 2593 | cerebral creatine deficiency syndrome 2 | AR |
| GBA | 2629 | gaucher_disease | AR |
| GBE1 | 2632 | glycogen_storage_disease | AR |
| GCDH | 2639 | glutaric_acidemia | AR |
| GFM1 | 85476 | combined_oxidative_phosphorylation_deficiency | AR |
| GJB1 | 2705 | charcot-marie-tooth_disease | XL |
| GJB2 | 2706 | non-syndromic hearing loss (GJB2-related) | AR |
| GLA | 2717 | fabry_disease | XL |
| GLB1 | 2720 | gm1_gangliosidosis | AR |
| GLDC | 2731 | glycine_encephalopathy | AR |
| GLE1 | 2733 | lethal_arthrogryposis_with_anterior_horn_cell_disease | AR |
| GNE | 10020 | inclusion_myopathy | AR |
| GNPTAB | 79158 | mucolipidosis | AR |
| GNPTG | 84572 | mucolipidosis | AR |
| GNS | 2799 | mucopolysaccharidosis | AR |
| GP1BA | 2811 | bernard-soulier_syndrome | AR |
| GP9 | 2815 | bernard-soulier_syndrome | AR |
| GPR56 | 9289 | bilateral_polymicrogyria | AR |
| GRHPR | 9380 | primary_hyperoxaluria | AR |
| HADHA | 3030 | mitochondrial_trifunctional_protein_deficiency | AR |
| HAX1 | 10456 | congenital_neutropenia | AR |
| HBA1 | 3039 | alpha-thalassemia | AR |
| HBA2 | 3040 | alpha-thalassemia | AR |
| HBB | 3043 | sickle_cell_anemia | AR |
| HEXA | 3073 | tay-sachs_disease | AR |
| HEXB | 3074 | sandhoff_disease | AR |
| HFE2 | 148738 | hemochromatosis | AR |
| HGSNAT | 138050 | mucopolysaccharidosis | AR |
| HLCS | 3141 | holocarboxylase_synthetase_deficiency | AR |
| HMGCL | 3155 | HMG-CoA lyase deficiency | AR |
| HOGA1 | 112817 | primary_hyperoxaluria | AR |
| HPS1 | 3257 | hermansky-pudlak_syndrome | AR |
| HPS3 | 84343 | hermansky-pudlak_syndrome | AR |
| HSD17B4 | 3295 | d-bifunctional_protein_deficiency | AR |
| HSD3B2 | 3284 | 3-beta-hydroxysteroid dehydrogenase type II deficiency | AR |
| HYAL1 | 3373 | mucopolysaccharidosis | AR |
| HYLS1 | 219844 | hydrolethalus_syndrome | AR |
| IDS | 3423 | mucopolysaccharidosis | XL |
| IDUA | 3425 | mucopolysaccharidosis | AR |
| IKBKAP | 8518 | familial dysautonomia | AR |
| IL2RG | 3561 | severe_combined_immunodeficiency | XL |
| IVD | 3712 | isovaleric_acidemia | AR |
| KCNJ11 | 3767 | familial_hyperinsulinism | AR |
| LAMA3 | 3909 | junctional_epidermolysis_bullosa | AR |
| LAMB3 | 3914 | junctional_epidermolysis_bullosa | AR |
| LAMC2 | 3918 | junctional_epidermolysis_bullosa | AR |
| LCA5 | 167691 | leber_congenital_amaurosis | AR |
| LDLR | 3949 | familial_hypercholesterolemia | AR |
| LDLRAP1 | 26119 | familial_hypercholesterolemia | AR |
| LHX3 | 8022 | combined_pituitary_hormone_deficiency | AR |
| LIFR | 3977 | stuve-wiedemann_syndrome | AR |
| LIPA | 3988 | wolman_disease | AR |
| LOXHD1 | 125336 | deafness,_autosomal_recessive | AR |
| LPL | 4023 | lipoprotein_lipase_deficiency | AR |
| LRPPRC | 10128 | leigh_syndrome | AR |
| MAN2B1 | 4125 | alpha-mannosidosis | AR |
| MCCC1 | 56922 | 3-methylcrotonyl-coa_carboxylase_deficiency | AR |
| MCCC2 | 64087 | 3-methylcrotonyl-coa_carboxylase_deficiency | AR |
| MCOLN1 | 57192 | mucolipidosis | AR |
| MED17 | 9440 | infantile_cerebral_and_cerebellar_atrophy | AR |
| MEFV | 4210 | familial_mediterranean_fever | AR |
| MESP2 | 145873 | spondylocostal_dysostosis | AR |
| MFSD8 | 256471 | neuronal_ceroid-lipofuscinosis | AR |
| MKS1 | 54903 | bardet-biedl_syndrome | AR |
| MLC1 | 23209 | megalencephalic_leukoencephalopathy_with_subcortical_cysts | AR |
| MMAA | 166785 | methylmalonic_acidemia | AR |
| MMAB | 326625 | methylmalonic_acidemia | AR |
| MMACHC | 25974 | methylmalonic_acidemia | AR |
| MMADHC | 27249 | methylmalonic_acidemia | AR |
| MPI | 4351 | congenital_disorder_of_glycosylation | AR |
| MPL | 4352 | congenital_amegakaryocytic_thrombocytopenia | AR |
| MPV17 | 4358 | mitochondrial_dna_depletion_syndrome | AR |
| MTHFR | 4524 | homocystinuria | AR |
| MTM1 | 4534 | x-linked_centrotubular_myopathy | XL |
| MTRR | 4552 | homocystinuria | AR |
| MTTP | 4547 | abetalipoproteinemia | AR |
| MUT | 4594 | methylmalonic_acidemia | AR |
| MYO7A | 4647 | usher_syndrome | AR |
| NAGLU | 4669 | mucopolysaccharidosis type IIIB | AR |
| NAGS | 162417 | n-acetylglutamate_synthase_deficiency | AR |
| NBN | 4683 | nijmegen_breakage_syndrome | AR |
| NDRG1 | 10397 | charcot-marie-tooth_disease | AR |
| NDUFAF5 | 79133 | mitochondrial_complex_i_deficiency | AR |
| NDUFS6 | 4726 | mitochondrial_complex_i_deficiency | AR |
| NEB | 4703 | nemaline_myopathy | AR |
| NPC1 | 4864 | niemann-pick_disease | AR |
| NPC2 | 10577 | niemann-pick_disease | AR |
| NPHS1 | 4868 | nephrotic_syndrome | AR |
| NPHS2 | 7827 | nephrotic_syndrome | AR |
| NR2E3 | 10002 | enhanced_s-cone_syndrome | AR |
| NTRK1 | 4914 | congenital insensitivity to pain with anhidrosis | AR |
| OAT | 4942 | ornithine_aminotransferase_deficiency | AR |
| OPA3 | 80207 | 3-methylglutaconic_aciduria | AR |
| OTC | 5009 | ornithine_carbamoyltransferase_deficiency | XL |
| PAH | 5053 | phenylketonuria | AR |
| PCCA | 5095 | propionic_acidemia | AR |
| PCCB | 5096 | propionic_acidemia | AR |
| PCDH15 | 65217 | usher_syndrome | AR |
| PDHA1 | 5160 | pyruvate_dehydrogenase_deficiency | XL |
| PDHB | 5162 | pyruvate_dehydrogenase_deficiency | AR |
| PEX1 | 5189 | zellweger_syndrome_spectrum | AR |
| PEX10 | 5192 | zellweger_syndrome_spectrum | AR |
| PEX2 | 5828 | zellweger_syndrome_spectrum | AR |
| PEX6 | 5190 | zellweger_syndrome_spectrum | AR |
| PEX7 | 5191 | rhizomelic_chondrodysplasia_punctata | AR |
| PFKM | 5213 | glycogen_storage_disease | AR |
| PHGDH | 26227 | 3-phosphoglycerate_dehydrogenase_deficiency | AR |
| PKHD1 | 5314 | polycystic_kidney_disease | AR |
| PMM2 | 5373 | congenital_disorder_of_glycosylation | AR |
| POMGNT1 | 55624 | muscular_dystrophy-dystroglycanopathy | AR |
| PPT1 | 5538 | neuronal_ceroid-lipofuscinosis | AR |
| PROP1 | 5626 | combined_pituitary_hormone_deficiency | AR |
| PRPS1 | 5631 | Charcot-Marie-Tooth disease, type 5 / Arts syndrome / deafness, X-linked 1 | XL |
| PSAP | 5660 | combined_prosaponin_deficiency | AR |
| PTS | 5805 | 6-pyruvoyl-tetrahydropterin synthase deficiency | AR |
| PUS1 | 80324 | mitochondrial_myopathy_and_sideroblastic_anemia | AR |
| PYGM | 5837 | glycogen_storage_disease | AR |
| RAB23 | 51715 | carpenter_syndrome | AR |
| RAG2 | 5897 | omenn_syndrome | AR |
| RAPSN | 5913 | congenital_myasthenic_syndrome | AR |
| RARS2 | 57038 | pontocerebellar_hypoplasia | AR |
| RDH12 | 145226 | leber_congenital_amaurosis | AR |
| RMRP | 6023 | cartilage-hair_hypoplasia | AR |
| RPE65 | 6121 | leber_congenital_amaurosis | AR |
| RPGRIP1L | 23322 | joubert_syndrome | AR |
| RS1 | 6247 | x-linked_juvenile_retinoschisis | XL |
| RTEL1 | 51750 | dyskeratosis_congenita | AR |
| SACS | 26278 | spastic_ataxia | AR |
| SAMHD1 | 25939 | aicardi-goutieres_syndrome | AR |
| SEPSECS | 51091 | pontocerebellar_hypoplasia | AR |
| SGCA | 6442 | limb-girdle_muscular_dystrophy | AR |
| SGCB | 6443 | limb-girdle_muscular_dystrophy | AR |
| SGCG | 6445 | limb-girdle_muscular_dystrophy | AR |
| SGSH | 6448 | mucopolysaccharidosis | AR |
| SLC12A3 | 6559 | gitelman_syndrome | AR |
| SLC12A6 | 9990 | andermann_syndrome | AR |
| SLC17A5 | 26503 | salla_disease | AR |
| SLC22A5 | 6584 | primary_carnitine_deficiency | AR |
| SLC25A13 | 10165 | citrullinemia | AR |
| SLC25A15 | 10166 | hyperornithinemia-hyperammonemia-homocitrullinuria_syndrome | AR |
| SLC26A2 | 1836 | sulfate transporter-related osteochondrodysplasia | AR |
| SLC26A4 | 5172 | pendred_syndrome | AR |
| SLC35A3 | 23443 | arthrogryposis,_mental_retardation,_and_seizures | AR |
| SLC37A4 | 2542 | glycogen_storage_disease | AR |
| SLC39A4 | 55630 | acrodermatitis_enteropathica | AR |
| SLC4A11 | 83959 | corneal_dystrophy_and_perceptive_deafness | AR |
| SLC6A8 | 6535 | creatine_transporter_deficiency | XL |
| SLC7A7 | 9056 | lysinuric_protein_intolerance | AR |
| SMARCAL1 | 50485 | schimke_immunoosseous_dysplasia | AR |
| SMN1 | 6606 | spinal_muscular_atrophy | AR |
| SMPD1 | 6609 | niemann-pick_disease | AR |
| STAR | 6770 | congenital_adrenal_hyperplasia | AR |
| SUMF1 | 285362 | multiple_sulfatase_deficiency | AR |
| TCIRG1 | 10312 | osteopetrosis,_autosomal_recessive | AR |
| TECPR2 | 9895 | hereditary_spastic_paraplegia | AR |
| TFR2 | 7036 | hemochromatosis | AR |
| TGM1 | 7051 | inherited_ichthyosis | AR |
| TH | 7054 | segawa_syndrome | AR |
| TMEM216 | 51259 | joubert_syndrome | AR |
| TPP1 | 1200 | neuronal_ceroid-lipofuscinosis | AR |
| TRMU | 55687 | infantile_liver_failure | AR |
| TSFM | 10102 | combined_oxidative_phosphorylation_deficiency | AR |
| TTPA | 7274 | ataxia_with_isolated_vitamin_e_deficiency | AR |
| TYMP | 1890 | mitochondrial_dna_depletion_syndrome | AR |
| USH1C | 10083 | usher_syndrome | AR |
| USH2A | 7399 | usher_syndrome | AR |
| VPS13A | 23230 | choreoacanthocytosis | AR |
| VPS13B | 157680 | cohen_syndrome | AR |
| VPS45 | 11311 | congenital_neutropenia | AR |
| VRK1 | 7443 | pontocerebellar_hypoplasia | AR |
| VSX2 | 338917 | isolated_anophthalmia-microphthalmia | AR |
| WNT10A | 80326 | odonto-onycho-dermal_dysplasia | AR |
