## Supplemental Figure 1 for "An algorithm to identify patients with rare genetic disorders and its real-world data application"

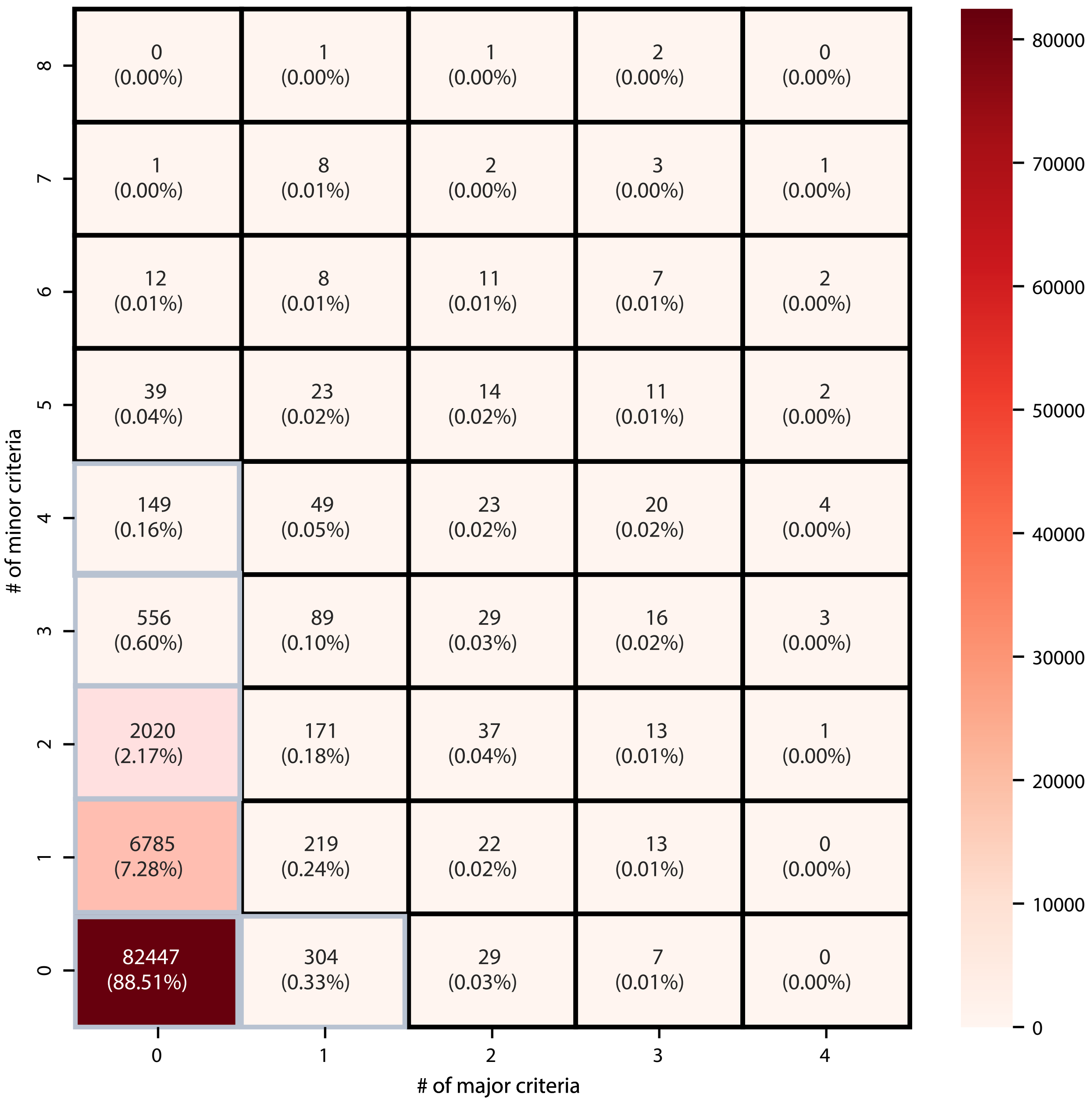


Supplementary Figure 1: Number of children with each number of major and minor criteria from PheIndex digital phenotype. Square with grey borders indicate major/minor criteria combination that result in a "positive" label from the digital phenotype.
