## Supplemental Figure 2 for "An algorithm to identify patients with rare genetic disorders and its real-world data application"

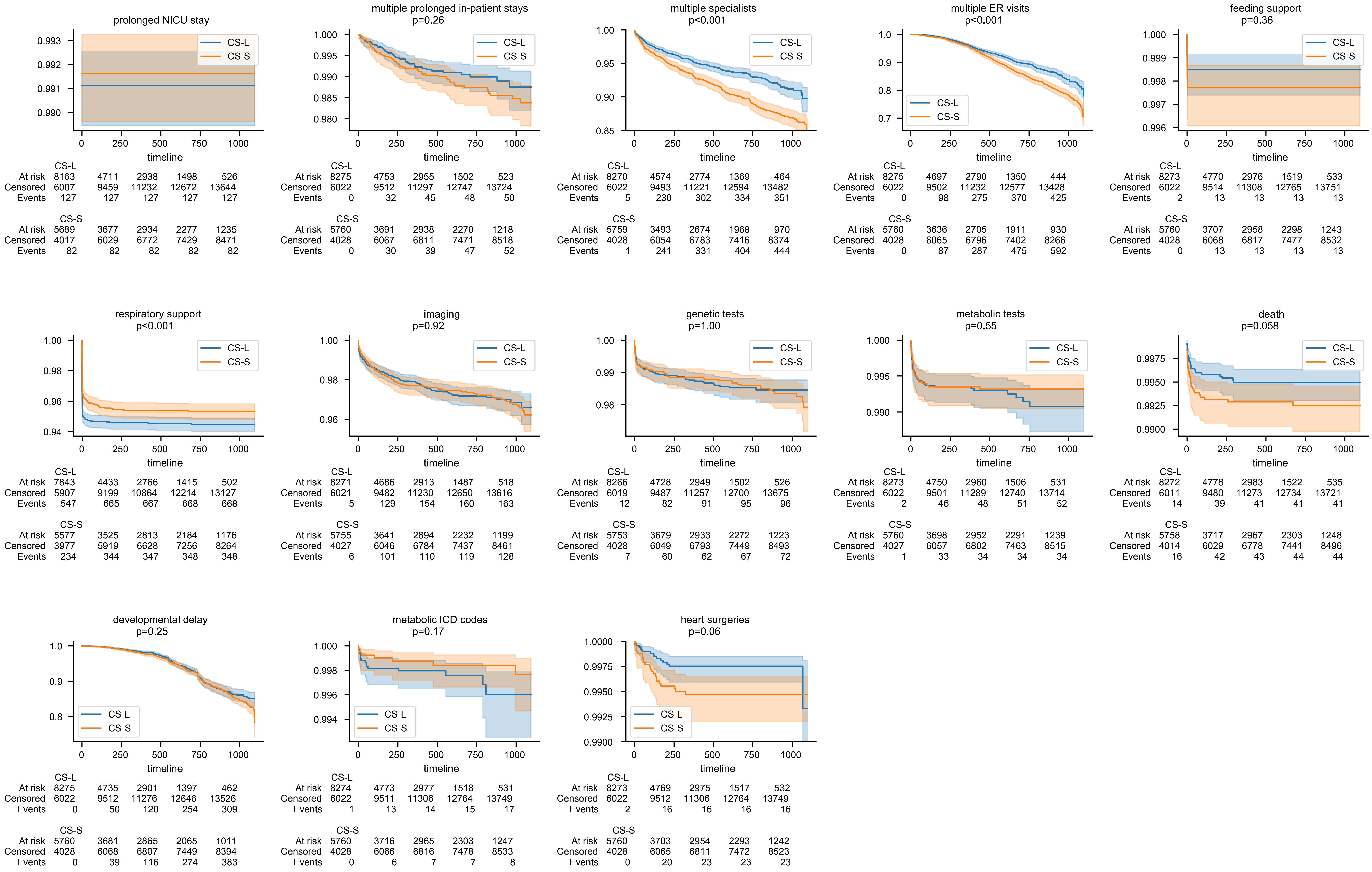


Supplementary Figure 2: Kaplan-Meier survival curves for each of the 13 PheIndex criteria for large panel carrier screening (CS-L, in blue) and small panel carrier screening (CS-S, in orange) cohorts. P-value from log-rank test bewteen CS-L and CS-S is reported under the title for an individual criteria.
