## Supplemental Figure 3 for "An algorithm to identify patients with rare genetic disorders and its real-world data application"

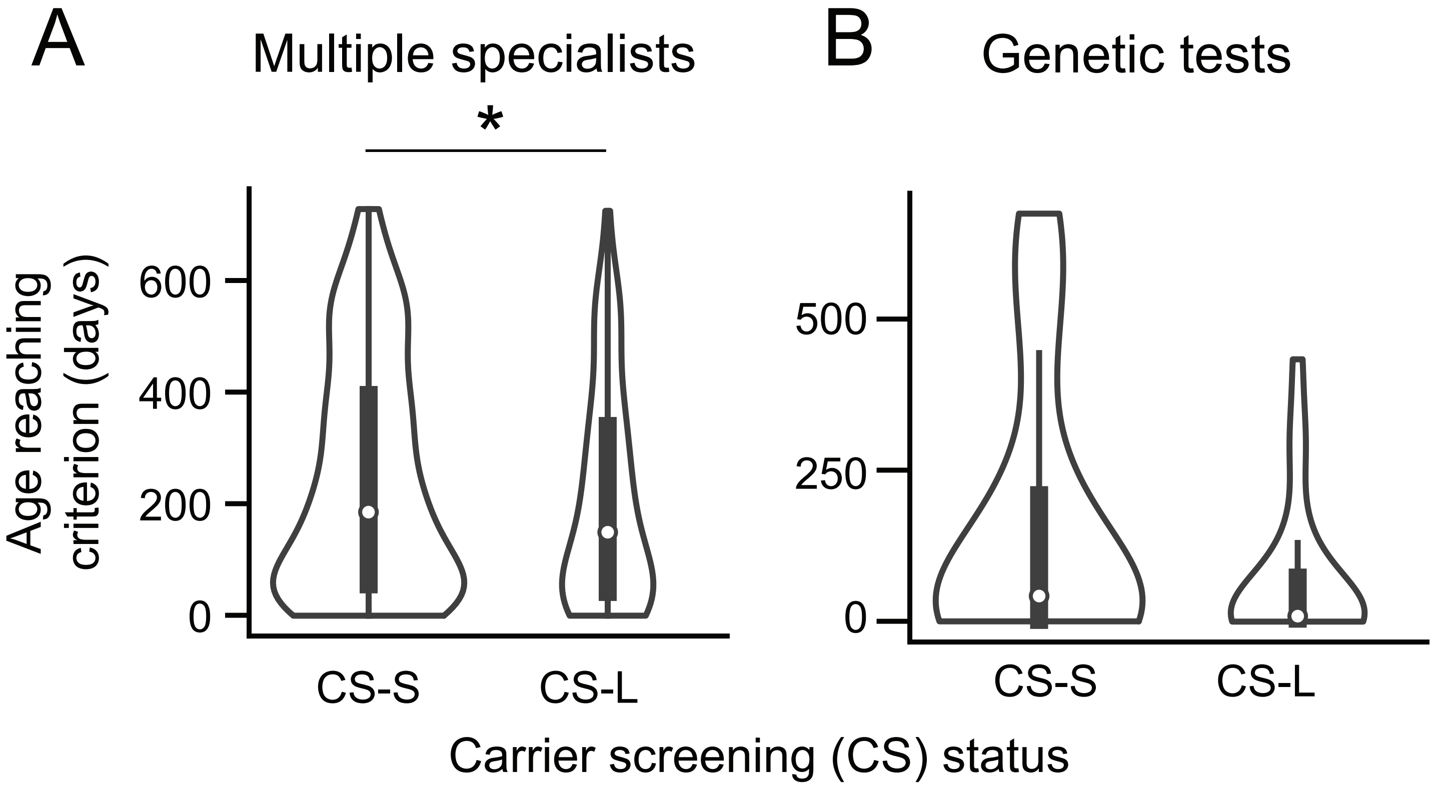


Figure S3: (A) Distribution of ages when the multiple specialists criterion is first met for children whose mothers received small panel / CS-S (median=192, n=331) and large panel / CS-L (median=148, n=158) testing. Only patients with two years of follow-up data are included, and only the first two years of clinical data are used for determining meeting criterion. * indicates a difference in distributions at signficance level p<0.05 (Mann-Whitney U-test).

(B) Distribution of ages when the genetic testing criterion is met for children whose mothers received CS-S (median=42, n=34) and CS-L (median=8.5, n=22) testing.
