## Supplemental Figure 4 for "An algorithm to identify patients with rare genetic disorders and its real-world data application"

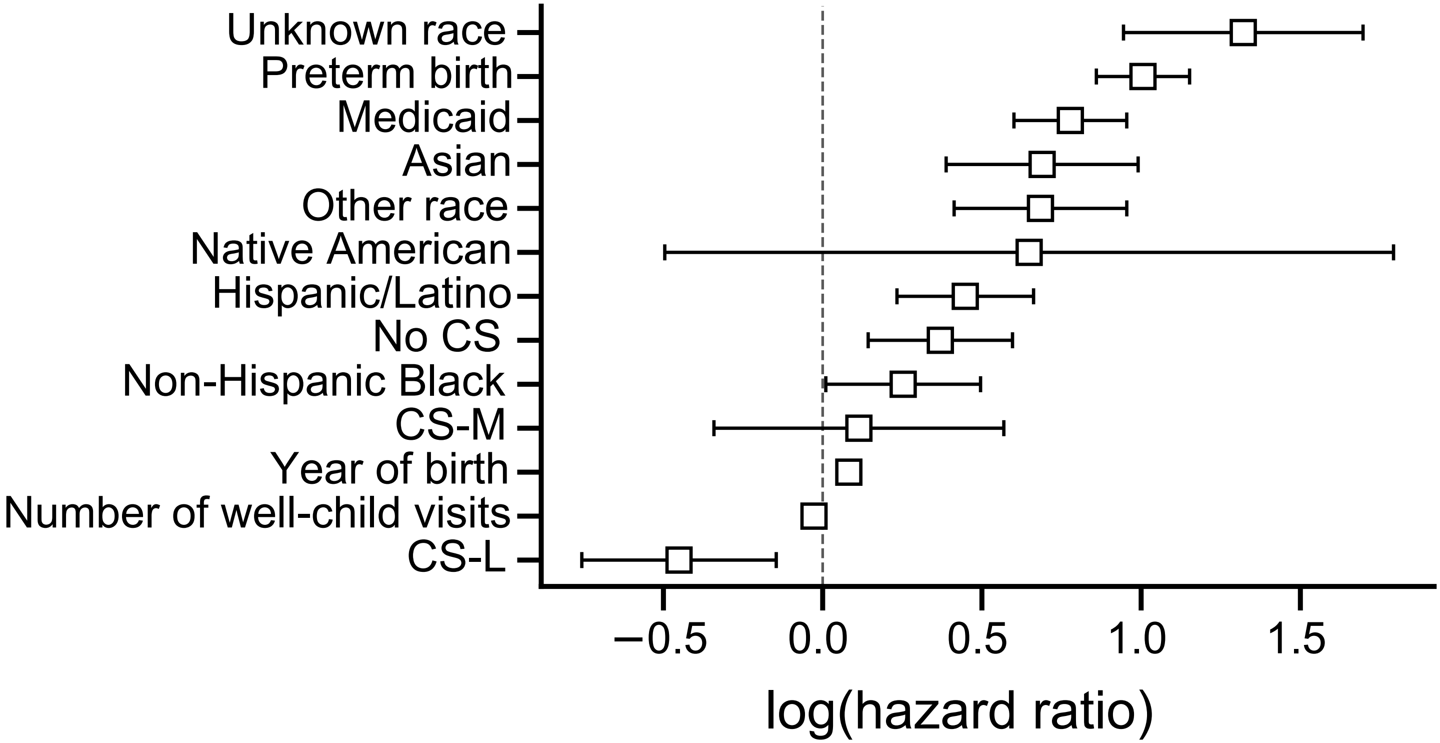


Figure S4: Coefficients from Cox proportional hazards model for PheIndex binary classification. Error bars show 95% confidence intervals. The exposure of interest is carrier screening (CS) status (No CS, CS-S/small, CS-M/medium, CS-L/large), where CS-S is the referent category. Confounders controlled for include: race/ethnicity (where Non-Hispanic White is the referent), preterm birth, health insurance (Medicaid vs not Medicaid), year of birth, and number of well-child visits.
